# COVID-19 causing HELLP-like syndrome in pregnancy and role of angiogenic factors for differential diagnosis

**DOI:** 10.1101/2020.07.10.20133801

**Authors:** Francesc Figueras, Elisa Llurba, Raigam Martinez-Portilla, Josefina Mora, Fatima Crispi, Eduard Gratacos

## Abstract

**Importance:** The clinical presentation of hemolysis, elevated liver enzymes, and low platelet count (HELLP) syndrome is one of the more severe forms of preeclampsia. COVID-19 infection exhibits signs that are shared with preeclampsia and HELLP syndrome, which may lead to needless interventions and iatrogenic preterm delivery.

**Objective:** We evaluated the prevalence of HELLP-like signs in pregnant women admitted for COVID-19 and the value of angiogenic factors to rule out preeclampsia.

**Methods:** a consecutive series of 27 pregnant women beyond 20 weeks of gestation, with symptomatic COVID-19. Clinical and analytical features were recorded and those cases with signs of HELLP syndrome were tested for sFlt-1/PlGF ratio.

**Results:** Seven patients (25.9%) presented at least one sign of suspected HELLP syndrome, of which 2 (7.4%) were diagnosed clinically with PE because of hypertension and high transaminases and 5 (18.5%) had only elevated transaminases. sFlt-1/PlGF ratio was normal in 6 of 7.

**Conclusion:** Symptomatic COVID-19 may simulate severe preeclampsia in pregnancy. Angiogenic factors may be essential to avoid false diagnosis and needless interventions.

**KEY-POINTS:** *Question:* Do pregnant women with symptomatic COVID-19 infection exhibit signs shared with preeclampsia and HELLP syndrome? Are these conditions ruled out by the measurement of angiogenic factors?

*Findings:* In series of 27 pregnancies with symptomatic COVID-19 infection, 7 presented at least one sign of suspected HELLP syndrome. In 6 of them, the level of angiogenic factors ruled out preeclampsia.

*Meaning:* Symptomatic COVID-19 may simulate severe pregnancy in pregnancy. Angiogenic factors may be essential to avoid false diagnosis and needless interventions. These data were presented in a Virtual Symposium on Covid-19 and Pregnancy on 17 April: 2020:(http://medicinafetalbarcelona.org/simposiocovid19/ [Spanish] and https://medicinafetalbarcelona.org/symposiumcovid19/ [English]

## Introduction

The COVID-19 outbreak has now rapidly spread worldwide, being declared by the WHO a pandemic infection on March 11^th^, with more than 1.2 million reported infections and 67,000 deaths (April 6^th^) (1). With more than 130,000 infections and 12,000 deaths, Spain is the second country in number of cases (1). The features of COVID-19 in pregnancy are still poorly described.

Symptomatic COVID-19 is characterized by flu-like symptoms, but in around 15% of cases it is developed a severe respiratory compromise or multi-organ failure (2). Abnormal aminotransferase and lactate dehydrogenase (LDH) levels and proteinuria are common features, not only in critically ill patients but also in milder forms of COVID-19 (3)(4). These signs could be clinically relevant in pregnant women. In pregnancy, the clinical presentation of hemolysis, elevated liver enzymes, and low platelet count (HELLP) syndrome is one of the more severe forms of preeclampsia because it has been associated with increased rates of maternal morbidity and mortality. HELLP syndrome may have an insidious and atypical onset, with up to 15% of the patients lacking either hypertension (5). Therefore, COVID-19 signs in pregnant women might lead to a misdiagnosis of preeclampsia and iatrogenic preterm delivery. Maternal levels of the angiogenic factors placental growth factor (PlGF), soluble fms-like tyrosine kinase, and particularly the ratio sFlt-1/PlGF have recently been proposed as highly specific markers in ruling out suspected preeclampsia (6).

There is no previous information on the frequency of preeclampsia-like signs in COVID-19 pregnant women and whether angiogenic factors might help in the differential diagnosis with true preeclampsia. In this series we recorded the clinical and analytical data of a consecutive series of 27 pregnant women with symptomatic COVID-19. We describe the prevalence of HELLP-like syndrome and the performance of sFlt-1/PlGF ratio to rule out true preeclampsia.

## Methods

A case-series was created of consecutive pregnant women of more than 20 weeks of gestation presenting with symptomatic COVID-19 between March 21^st^ and April 7^th^ 2020, at two hospitals in Barcelona, with symptomatic and confirmed COVID-19 infection by quantitative RT-PCR on nasopharyngeal swabs.

This study was approved by the Ethics Committee of Hospital Sant Joan de Deu (PIC-56-20).

Symptomatic disease was defined by the presence acute respiratory tract infection (sudden onset of at least one of the following: cough, fever [central temperature >38ºC], shortness of breath) with no other etiology that fully explains the clinical presentation

Management of patients followed the WHO guidance (7).

In women in which there was a clinical suspicion of HELLP syndrome, maternal serum levels of soluble fms-like tyrosine kinase 1 [sFlt-1] and placental growth factor [PlGF] and sFlt-1/PlGF ratio were determined by the fully automated Elecsys assays for sFlt-1 and PlGF on an electrochemiluminescence immunoassay platform (Cobas analyzers, Roche Diagnostics). Preeclampsia was defined according to ACOG criteria (5) and abnormal aminotransferase or lactate dehydrogenase levels were defined when above twice the upper laboratory reference value.

## Results

A total of 27 pregnant women were included. Table 1 shows the baseline characteristics of the population. Overall, 7 patients (25.9%) presented at least one sign of suspected HELLP syndrome, of which 2 (7.4%) were diagnosed clinically with PE because of hypertension and proteinuria or elevated transaminases; 3 (11.1%) had significant proteinuria plus elevated transaminases; and, 2 had isolated abnormal transaminase levels. sFlt-1/PlGF ratio was elevated over the cut-off for preeclampsia in 1 of 7. Of note, magnesium sulphate was initiated in 2 patients, and discontinued in those with normal levels. There were no other obstetrical complications. Table 2 shows the clinical characteristics of these seven pregnancies.

**Table 1.** Baseline characteristics of the included population

|  |  |
| --- | --- |
| Maternal age (years) | 33 (8);20-45 |
| Gestational age at admission (weeks) | 33.6 (8.2); 21.3-41.3 |
| Clinical symptoms |  |
| Cough | 20 (74.1) |
| Fever (>38°) | 15 (55.6) |
| Shortness of breath | 11 (40.7) |
| Diarrhea | 3 (11.1) |
| Respiratory rate (breaths/minute) | 22 (9);18-36 |
| Basal Oxygen Saturation (%) | 97 (5); 93-99 |
| Lymphocyte count (cells/ μL) | 1267 (760); 750-2130 |
| C-reactive protein (mg/dL) | 3.6 (6.8); 0.1-26 |
| AST (U/L) | 45 (50); 14-706 |
| ALT (U/L) | 37 (167); 11-363 |
| LDH (U/L) | 247 (75); 174-985 |
| D-Dimer (ng/mL) | 2474(2039); 904-4310 |
| Ongoing pregnancies | 16 (59.3) |
| Maternal complication | 0 |
| Oxygen support |  |
| Nasal cannula-Venturi mask | 15 (55.6) |
| CPAP | 1 (3.7) |
| Intubation | 0 |
| Hydroxichloroquine sulphate | 13 (48.1) |
| Azithromycin | 13 (48.1) |
| Cesarean section+ | 5 (45.6) |
| Birthweight (g)+ | 3300(574); 2566-3590 |
| Preterm birth (<37 weeks)+ | 0 |
| SGA (<10<sup>th</sup> centile) | 1 (3.7) |
| Neonatal complication+ | 0 |

Median (IQR) and range or n (%), as appropriate
+ n=12
IQR: Interquartile range; CPAP: Continuous Positive Airway Pressure; AST: Aspartate aminotransferase; ALT: Alanine aminotransferase; LDH: Lactate dehydrogenase; SGA: Small-for-Gestational Age

**Table 2.** Clinical characteristics of women with HELLP syndrome or preeclampsia suspicion

|  | Case 1 | Case 2 | Case 3 | Case 4 | Case 5 | Case 6 | Case 7 |
| --- | --- | --- | --- | --- | --- | --- | --- |
| Maternal age (years) | 33 | 24 | 29 | 36 | 32 | 43 | 26 |
| Gestational age at admission (weeks) | 30<sup>+5</sup> | 37<sup>+2</sup> | 37<sup>+0</sup> | 33<sup>+4</sup> | 33<sup>+6</sup> | 21<sup>+3</sup> | 33<sup>+0</sup> |
| Respiratory rate (breaths/minute) | 25 | 18 | 30 | 28 | 34 | 14 | 23 |
| Basal Oxygen Saturation (%) | 98 | 98 | 94 | 98 | 93 | 99 | 97 |
| Lymphocyte count (x10<sup>9</sup>/L) | 2 | 1.6 | 1.49 | 2.4 | 1.3 | 1.5 | 1.7 |
| C-reactive protein (mg/dL) | NO |  | 13.24 | 30 | 4.8 | 22 | 45 |
| Platelet count (x10<sup>9</sup>/L) | 341 | 189 | 167 | 182 | 126 | 315 | 201 |
| Systolic blood pressure (mmHg) | 153 | 146 | 117 | 113 | 115 | 123 | 100 |
| Diastolic blood pressure (mmHg) | 83 | 84 | 65 | 73 | 69 | 58 | 55 |
| Proteinuria (mg/24h or dipstick) | 800 | - | 1178 | - | 602 | 1+ | 2+ |
| AST (U/L) | 146 | 76 | 706 | 156 | 175 | 177 | 77 |
| ALT (U/L) | 268 | 46 | 271 | 190 | 363 | 118 | 81 |
| LDH (U/L) | 227 | 360 | 985 | 301 | 282 | 345 | 243 |
| D-Dimer (mg/mL) | - | 2450 | 2300 | 1561 | 4020 | - | - |
| PlGF (pg/mL) | 506 | 257 | 77 | 141 | 206 | 302 | 625 |
| sFlt-1 (pg/mL) | 1385 | 21689 | 32375 | 9273 | 5119 | 3207 | 3098 |
| sFlt-1/PlGF ratio | 3 | 84 | 423 | 66 | 25 | 11 | 5 |
| Maternal complication | No | No | No | No | No | No | No |
| Gestational age at delivery | - | 37+2 | 37+0 | - | - | - | - |
| Birthweight | - | 2898 | 3300 | - | - | - | - |
| Neonatal RT-PCR | - | Negative | Negative | - | - | - | - |
| Neonatal complication | - | No | No | - | - | - | - |

AST: Aspartate aminotransferase; ALT: Alanine aminotransferase; LDH: Lactate dehydrogenase; PLGF: placental growth factor; sFlt-1: Soluble fms-like tyrosine kinase-1; RT-PCR: Real-Time Polymerase Chain Reaction

## Discussion

The effect of COVID-19 infection on pregnancy is not well known because of the lack of reliable data. Small series from China suggest that the clinical characteristics of COVID-19 in pregnancy do not differ from those reported in non-pregnant adults (8)(9)(10). In this series, rather than focusing on perinatal outcomes, we report that 26% of pregnant women presented clinical or analytical features currently considered for the diagnosis of preeclampsia or HELLP syndrome.

As already described in adults (3), we found abnormal aminotransferase or lactate dehydrogenase levels in one-third of cases. Massive proteinuria has also been reported in one-third of adults with COVID-19 (4). These signs are shared with preeclampsia, one of the most feared complications of pregnancy, which complicates 2% to 8% and is a leading cause of maternal morbidity and mortality (11) and iatrogenic prematurity (12). Accordingly, best practice guidelines for severe PE entails early treatment with magnesium sulphate and elective preterm delivery (5). Thus, obstetricians should be aware of the risk of misdiagnosing severe preeclampsia in COVID-19.

Other obstetric and medical disorders share clinical and laboratory findings with severe preeclampsia (13). Maternal levels of sFlt-1/PlGF, as markers of placental vasculogenesis, have a high performance in ruling out suspicion of preeclampsia (6). This preliminary series suggest that the performance of angiogenic factors is high in women with COVID-19 infection, and thus measuring angiogenic factors seems critical in COVID-19 pregnancies with PE-like analytical signs to rule out true PE, avoiding needless interventions and iatrogenic preterm delivery.

## Data Availability

Data referred to in the manuscript are available on personal demand

